# Exponential increase in neutralizing and spike specific antibodies following vaccination of COVID-19 convalescent plasma donors

**DOI:** 10.1101/2021.02.02.21250836

**Authors:** Molly A. Vickers, Alan Sariol, Judith Leon, Alexandra Ehlers, Aaron V. Locher, Kerry A. Dubay, Laura Collins, Dena Voss, Abby E. Odle, Myrl Holida, Anna E. Merrill, Stanley Perlman, C. Michael Knudson

## Abstract

**Background:** With the recent approval of COVID-19 vaccines, recovered COVID-19 subjects who are vaccinated may be ideal candidates to donate COVID-19 convalescent plasma (CCP).

**Case Series:** Three recovered COVID-19 patients were screened to donate CCP. All had molecularly confirmed COVID-19, and all were antibody positive by chemiluminescence immunoassay (DiaSorin) prior to vaccination. All were tested again for antibodies 11 to 21 days after they received the first dose of the vaccine (Pfizer). All showed dramatic increases (∼50 fold) in spike-specific antibody levels and had at least a 20-fold increase in the IC50 neutralizing antibody titer based on plaque reduction neutralization testing (PRNT). The spike-specific antibody levels following vaccination were significantly higher than those seen in any non-vaccinated COVID-19 subjects tested to date at our facility.

**Conclusion:** Spike-specific and neutralizing antibodies demonstrated dramatic increases following a single vaccination post COVID-19 infection which significantly exceeded values seen with COVID-19 infection alone. Recovered COVID-19 subjects who are vaccinated may make ideal candidates for CCP donation.

## Introduction

The emergence of SARS-CoV-2, the cause of COVID-19, has resulted in intense efforts to identify new and effective treatments. The lack of proven effective anti-viral therapies against coronaviruses has led to the broad utilization of COVID-19 convalescent plasma (CCP) obtained from survivors of COVID-19 to treat patients with active disease^1-3^. While the mechanism of action of CCP is uncertain, the most prevalent hypothesis is that CCP contains neutralizing antibodies that limit viral spread and replication^4^. Multiple reports describe the rationale for this therapy and provide some evidence of efficacy ^5-10^. One recent study demonstrated that CCP reduced severe disease by nearly 50% when used within 72 hours of disease onset^10^. However, other studies have failed to show clear evidence of efficacy^8,11^. One study linked high titer CCP to better outcomes than low titer CCP^12^. While there are many possible explanations for the variable results with CCP to date, one potential explanation is that studies that employ just a single unit of CCP fails to provide enough dose to consistently improve outcomes. Transfusion of only one or two units would be expected to only modestly increase antibody levels in COVID-19 patients. Approaches to identify CCP donors with super physiological levels of antibodies may allow for future studies that would allow investigators to address these important and unanswered questions. With COVID-19 vaccine use increasing, there will likely be numerous vaccinated subjects who have recovered from COVID-19 and are able to donate CCP.

A CCP donor program was established in our academic hospital with an in-house donor center. This program was established under in IRB-approved protocol that obtained informed consent from all subjects allowing for COVID-19 related research on blood samples obtained from these subjects^13^. A biorepository of serum samples from these subjects was established that included serum samples from screening or CCP donation. Of these subjects, several were frontline workers and per hospital policy were offered COVID-19 vaccine if they were 90 or more days out from the infection. Given the uncertainty about the durability of the immune response in recovered COVID-19 patients, several of the subjects in our study elected to get the vaccine when offered and blood samples were obtained from them as part of this study. The eligibility of vaccinated subjects to donate convalescent plasma has been hotly debated by the convalescent plasma community with some believing that recovered COVID-19 subjects who are vaccinated may be ideal candidates for donation. The results described here examine antibody levels in these subjects and compares these levels to those detected following infection.

## Materials and Methods

### CCP donor screening and testing

Potential CCP donors were screened following FDA guidance instructions under an IRB approved protocol (#202003554). The consent signed by all subjects allowed the use of blood samples for research purposes. After vaccine administration was initiated at our facility, subjects were contacted to obtain additional blood samples for research as allowed under this protocol. The date of diagnosis was recorded for all subjects based on positive molecular or antibody testing of the subject. Since the date if disease onset was not systematically recorded for these subjects, the date of diagnosis is defined as day 0 for the purposes of this study. In subjects with molecular testing, this was generally within a week of symptom onset. Serum from all subjects was stored at -30C or colder prior to SARS-CoV-2 antibody testing.

### SARS-CoV-2 immunoassays

Samples for this study were tested using the DiaSorin LIAISON® SARS-CoV-2 S1/S2 IgG chemiluminescence immunoassay (Saluggia, Italy) which has a positive cutoff of 15 arbitrary units per ml (AU/ml) and an upper range limit of 400 AU/ml ^14^. Samples above 400 AU/ml were diluted with antibody negative serum until the value was within the assay range. For these samples, the value reported represents the measured value multiplied by the fold dilution. For example, if a sample was originally >400 AU/ml and a dilution of 100 ul subject serum into 400 ul of negative serum resulted in a value at 300 AU/ml the reported antibody level for that subject would be 1500 AU/ml (5 x 300). Some samples were also tested with the Roche Diagnostics Elecys Anti-SARS-CoV-2 electrochemiluminescence immunoassay, which target total antibodies (IgG, IgM, IgA) to the nucleocapsid protein. This assay uses a cut-off index (COI) of 1.0 or higher to indicate a positive result. Both the Roche and the DiaSorin assays were granted emergency use authorization by the FDA and demonstrate similar performance^14^.

### SARS-CoV-2 Neutralization assay

Vero E6 cells (available from ATCC) were maintained in 10% FBS-DMEM in a humidified incubator at 37°C in 5% CO_2_. The SARS-CoV-2 isolate 2019n-COV/USA-WA1/2019 was used for these studies (accession #MT123290). Plaque reduction assays were performed as previously described ^15^. Cultures were maintained for 3 days then fixed with 10% formaldehyde and stained with crystal violet for plaque quantification. The IC50 value was the calculated titer that would give a 50% reduction in the average number of plaques per plate^16^.

## Case Series

### Case 1

A male in his 60s was diagnosed and recovered from COVID-19 in the fall of 2020. His diagnosis was molecularly confirmed via a nasal swab and PCR testing on day 0, 2 days after he developed symptoms consistent with COVID-19. He recovered without hospitalization or treatment by day 12 and consented to participate in our CCP study on day 70. Antibody screening using the DiaSorin immunoassay was positive (109 AU/ml) and above our cutoff of 100 AU/ml so he successfully donated CCP on day 76. At that time his SARS-CoV2 IgG antibody level was below our cutoff so he was not asked to donate CCP again. He received the Pfizer vaccine on day 103 and experienced arm pain, body aches and a headache, all symptoms commonly observed with vaccination. He returned to donate platelets on day 120 when additional research samples were obtained for SARS-CoV-2 specific (immunoassay and neutralizing) antibody measurements. These were compared to samples obtained prior to his vaccination (Table 1). His antibody level by immunoassay was estimated to be 3940 AU/mL, nearly forty-fold higher than it was prior to vaccination. The PRNT assay demonstrated that his IC50 neutralizing antibody titer on day 76 was 286 while on day 120, 17 days after his first vaccination, the IC50 neutralizing antibody titer was 6753. This represents a 24-fold increase in his neutralizing antibody titer. The spike immunoassay was repeated 14 days after his second vaccination and the level was 4137 AU/ml, roughly the same as after his first vaccination. The timeline for the disease, screening, donations and vaccinations for all 3 subjects are shown in **Figure 1**.

**Table 1.** Vaccinated subject antibody levels.

| **Table 1. Vaccinated subject antibody levels** |  | **Pre** | **Post** | **Fold Change** |
| --- | --- | --- | --- | --- |
| **Case 1**<br>**M-P** | Spike IgG (AU/ml) | **98.7** | **3940** | **40** |
|  | Nucleocapsid Total Ig (COI) | **165** | **180** | **1.1** |
|  | PRNT IC50 titer | **286** | **6753** | **24** |
| **Case 2**<br>**F-P** | Spike IgG (AU/ml) | **24.1** | **1515** | **63** |
|  | Nucleocapsid Total Ig (COI) | **40.8** | **25.6** | **0.6** |
|  | PRNT IC50 titer | **<20** | **2350** | **>117** |
| **Case 3**<br>**M-P** | Spike IgG (AU/ml) | **63.6** | **2960** | **47** |
|  | Nucleocapsid Total Ig (COI) | **3.42** | **1.82** | **0.5** |
|  | PRNT IC50 titer | **652** | **>14580** | **>22** |

**Figure 1:**
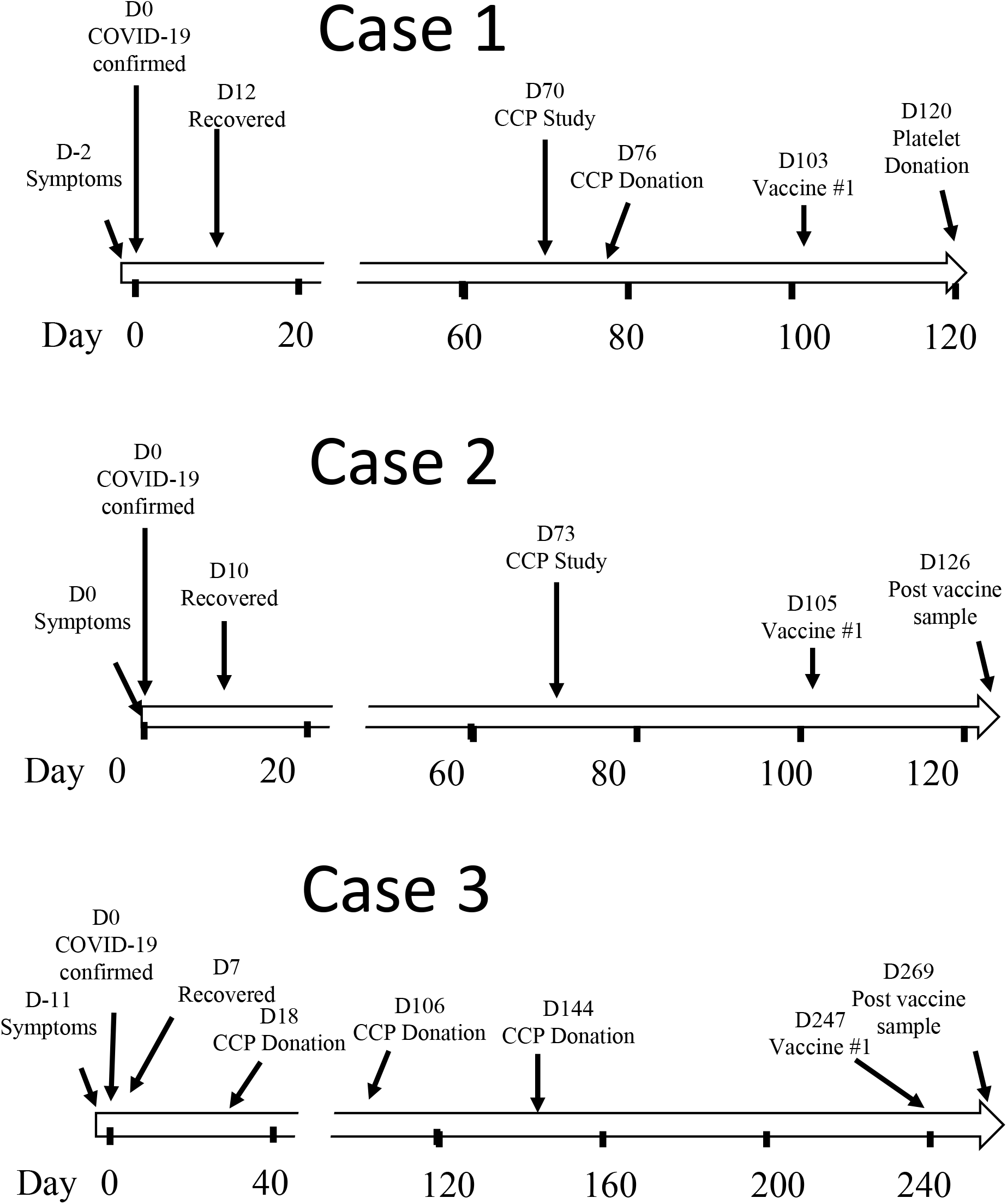
Subject timelines. The timeline for the three primary CCP subjects described in this report. Day 0 is the day that COVID-19 was confirmed by nasal swab molecular testing.

### Case 2

A female in her 60s was diagnosed and recovered from COVID-19 in the fall of 2020. Her diagnosis was molecularly confirmed via a nasal swab and PCR testing the same day she developed symptoms consistent with COVID-19. She recovered without hospitalization or treatment by day 10 and consented to participate in our CCP study on day 70. Antibody screening using the DiaSorin immunoassay was positive (24.1 AU/ml), but below our cutoff for donation, so she did not donate CCP. She received the Pfizer vaccine on day 105 and returned when additional research samples were obtained on day 126 for SARS-CoV-2 specific antibody testing (Table 1). Her antibody level by immunoassay was estimated to be 1515 AU/ml, about a sixty-fold increase relative to her antibody level 35 days prior to vaccination. The PRNT assay demonstrated that her IC50 neutralizing antibody titer on day 70 was <20 while on day 126, 21 days after her first vaccination, the IC50 neutralizing antibody titer was 2350. This represents a >117-fold increase in her neutralizing antibody titer. The spike immunoassay was repeated 14 days after her second vaccination and the level was 1452 AU/ml, roughly the same as after the first vaccination.

### Case 3

A male in his 50s was diagnosed and recovered from COVID-19 in the spring of 2020. His diagnosis was molecularly confirmed via a nasal swab and PCR testing on day 0, 11 days after he developed symptoms consistent with COVID-19. He recovered without hospitalization or treatment by day 7 and consented to participate in our CCP study on day 14. Antibody screening using the DiaSorin immunoassay was positive (55.2 AU/ml) and he successfully donated CCP on day 18, 119 and 147. The cutoff for CCP donation was increased to 100 AU/ml after that and he was no longer eligible to donate CCP. He received the Pfizer vaccine on day 247 and returned when additional research samples were obtained on day 269, 22 days after he received the first vaccine. These were compared to samples obtained prior to his vaccination on day 147 (**Table 1**). His antibody level by immunoassay was estimated to be 2960 AU/ml, more than forty-fold higher than it was prior to vaccination. The PRNT assay demonstrated that his IC50 neutralizing antibody titer on day 147 was 652 while on day 269, 22 days after his first vaccination, the IC50 neutralizing antibody titer was >14589. This represents a 22-fold increase in his neutralizing antibody titer. The spike immunoassay was repeated 14 days after his second vaccination and the level was 3410 AU/ml, just slightly higher than after the first vaccination.

### ELISA Results on COVID-19 infected subjects

During this study, 126 subjects with molecularly confirmed COVID-19 have been tested using the DiaSorin immunoassay. The initial testing results on these subjects is shown in **Figure 2A**. As previously reported for other populations^14^, a relatively high (21%) fraction of COVID-19 patients were found to be antibody negative (<15 AU/ml). A majority (57%) were positive but had values under 100 AU/ml. Only 22% had values over 100 AU/ml which is the antibody level currently in use for CCP donation eligibility at our site. The mean of the initial positive results was 81.7 AU/ml and range was 15-324 AU/ml. Looking at all test results from this study, the DiaSorin immunoassay has been positive on 205 samples from 145 subjects in this study. When we look at these positive results relative to when subjects were confirmed to have COVID-19 (initial molecular or serological testing), all but one was under 500 AU/ml. This contrast sharply with the 3 cases detailed in this report and shown as solid triangles in **Figure 2B**.

**Figure 2:**
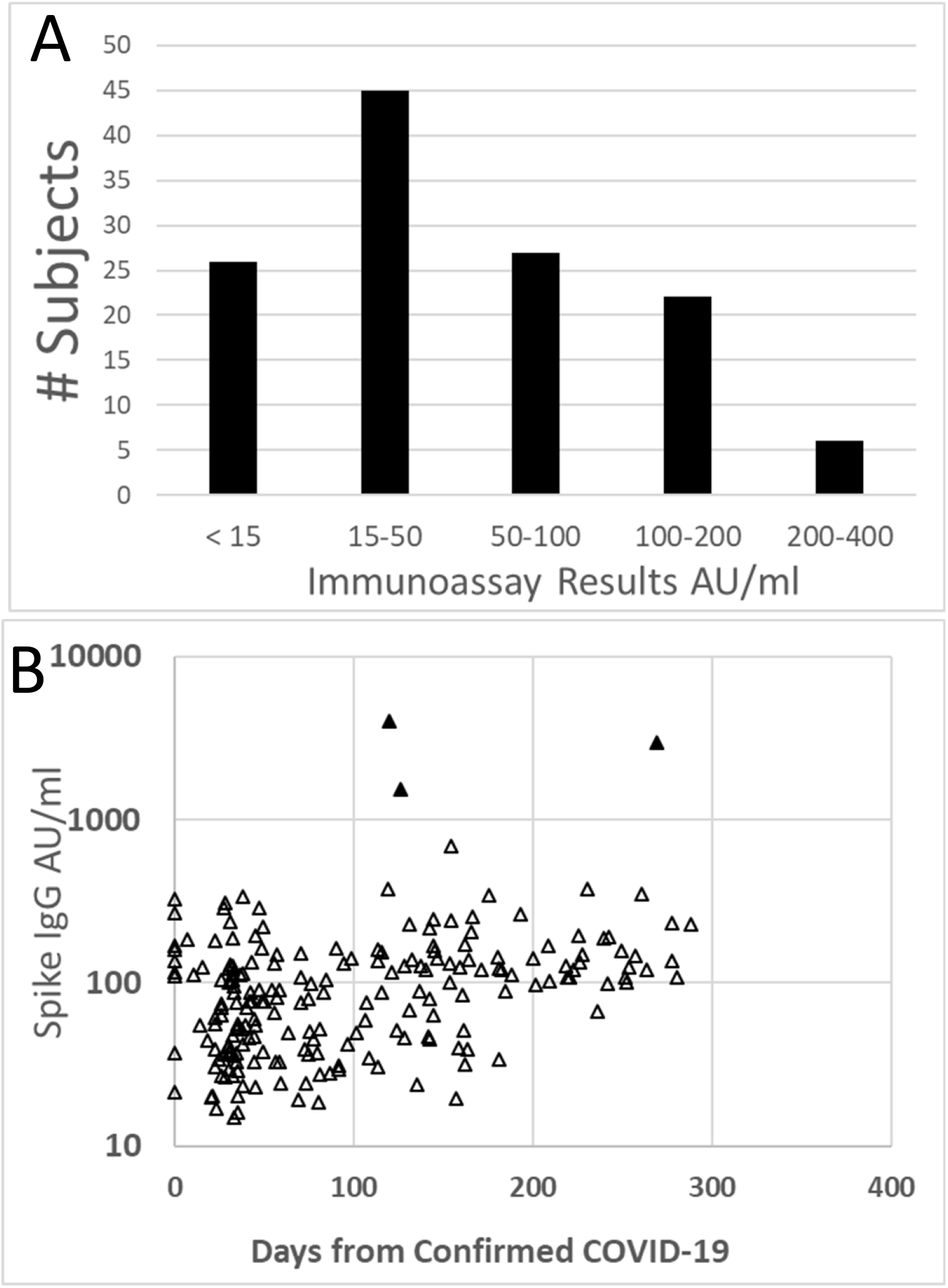
Spike specific IgG levels from CCP subjects. A) The initial/screening immunoassay results (DiaSorin) on 126 subjects with molecularly confirmed COVID-19 is shown. Of the positive results, the mean was 81 (Range 15-324). B) The spike specific immunoassay results for all positive<u>(>15)</u>samples are shown relative to the time the subject was confirmed to have COVID-19. The open triangles are non-vaccinated subjects and the filled triangles are from the 3 vaccinated cases outlined in this report.

### SPIKE ELISA levels on additional subjects

Given the dramatic increases seen in the 3 subjects described here, 8 additional CCP subjects were recruited following vaccination. All 8 show significant increases in the spike IgG levels as measured by immunoassay (**Table 2**). Two of these subjects received the Moderna (M) vaccine and both showed a significant increase in antibody levels suggesting that dramatic increases in antibody levels are not unique to the Pfizer vaccine. Of note, only one subject in this cohort had antibody levels less than 1000 AU/ml and this subject was the only subject tested to date who was antibody negative (<3.8 AU/ml) at the time they were screened. This suggests that this subject did not mount an antibody response following COVID-19 infection and thus may have responded to the vaccine like an uninfected subject. Excluding this “antibody-negative” subject, the other 10 subjects had an average spike-specific IgG level of 4166 AU/ml (range 1235-7854). This compares to an average of 81.7 AU/ml in the non-vaccinated antibody positive subjects screened for this study (N=109).

**Table 2:** DiaSorin antibody levels on additional subjects who received either the Pfizer (P) or Moderna (M) vaccine.

| Age-Sex-Vaccine | Pre<br>(days before<br>vaccine) | 1<sup>st</sup> Vaccine Days<br>from diagnosis | Post<br>(days after<br>vaccine(s)) | Fold Change |
| --- | --- | --- | --- | --- |
| **60s-F-P** | **31.9**<br>**(23)** | **185** | **1235**<br>**(22)** | **39** |
| **50s-F-P** | **<3.8**<br>**(55)** | **113** | **245**<br>**(36/14)** | **66** |
| **40s-F-M** | **39.4**<br>**(254)** | **276** | **6132**<br>**(25)** | **156** |
| **40s-F-P** | **39.1**<br>**(118)** | **281** | **6027**<br>**(20)** | **154** |
| **30s-F-P** | **39.2**<br>**(247)** | **273** | **4851**<br>**(20)** | **124** |
| **20s-F-P** | **379**<br>**(20)** | **250** | **3810**<br>**(16)** | **10** |
| **40s-F-M** | **84**<br>**(123)** | **292** | **7854**<br>**(17)** | **93** |
| **60s-F-P** | **37.6**<br>**(146)** | **195** | **3333**<br>**(21)** | **89** |

## Discussion

The results described here in a total of 11 subjects suggest that antibody positive recovered COVID-19 patients mount a strong amnestic response following just a single vaccination with either the Pfizer or the Moderna vaccine. These subjects all had spike-specific antibody levels at least 10-fold higher than what was detected prior to vaccination and all subjects who were antibody positive values prior to vaccination had antibody levels greater than 1200 AU/ml using a spike-specific IgG immunoassay. This contrast with non-vaccinated subjects who had an average of 81.7 AU/ml in this study. These dramatic results suggest that recovered COVID-19 patients who were antibody-positive following the initial infection would be strong candidates for CCP donation. Whether CCP collected from these donors would be more efficacious than CCP from non-vaccinated subjects is not known but the recent demonstration that patients receiving high titer plasma may do better than other CCP recipients certainly supports this possibility^12^. The nearly 10-fold increase in antibody levels raises the possibility that one unit of plasma from these donors would be enough to increase antibody levels in recipients to levels seen following infection alone. We are currently planning to examine this question at our facility.

This study also provides data regarding the durability of an amnestic response in subjects who have recovered from COVID-19. The ten subjects in this study who were antibody-positive when initially screened for CCP donation were vaccinated from 103 to 292 days after they were diagnosed. Six subjects were vaccinated 8 months or more after there were diagnosed and all 6 had antibody levels greater than 2900 AU/ml after just a single vaccine. Given that a single dose of the Pfizer vaccine had antibody responses that were less than observed in recovered COVID-19 patients^17,18^, these results suggest that these subjects had a strong amnestic response elicited by the vaccine that was maintained for at least 8 months. The FDA has recently (Jan 15, 2021) issued a guidance for industry document on the collection of CCP that includes information about the use of CCP from vaccinated subjects. These guidelines state CCP can be collected from vaccinated subjects if the subject previously had confirmed COVID-19 and the CCP is collected within 6 months of the end of symptoms. It is unfortunate that under these guidelines, 8 of the 11 subjects in this study would not be eligible to donate CCP as they had recovered from COVID-19 more than 6 months before they were tested. Each of these subjects had antibody spike specific IgG levels higher than any of 100+ previous donors we have screened for this study and each of the 3 tested using the PRNT assay had extremely high levels of neutralizing antibody as well.

The COVID-19 vaccine trials (NCT04368728/Pfizer;NCT04470427/Moderna) excluded subjects who had recovered from COVID-19 so data regarding the use of these vaccines in these patients is lacking^19^. Given that limitation, public health officials have been left to develop their own policies regarding whether and/or when to vaccinate recovered COVID-19 patients. As mentioned earlier, our facility selected 90 days post infection before recovered COVID-19 patients are offered the vaccine. Other sites have shorter time periods as some studies have shown that antibody responses may wane following infection^20^. This small case series provides evidence that some, if not most, recovered patients have evidence of an amnestic antibody response 8-10 months following infection. In this case series, all (10 of 10) subjects who had been antibody positive prior to CCP donation had robust antibody response to just a single vaccination. Given that vaccine demand currently greatly exceeds supply, these results raise the potential that antibody testing could be used to determine vaccine eligibility. One possible scenario is that antibody negative patients could be eligible for the vaccine at any point after infection while antibody positives be deferred for longer periods of time. Additionally, the strong antibody responses in these subjects following the first vaccination suggests that the second vaccine dose may not be necessary. The relative stable antibody levels in our 3 subjects seen 2 weeks after the second vaccine seems to support this possibility.

In conclusion, this small case series provides evidence to support a strong amnestic antibody response in recovered COVID-19 subjects who had previously been antibody positive. These subjects may be preferentially selected for CCP donation and raises the possibility that CCP collected from these vaccinated “super donors” could be more efficacious to infected patients than the CCP that has been used to date.

## Data Availability

All de-identified data described in this manuscript is available if needed.

## Acknowledgements

We would like to thank staff at the state hygienic lab who assisted in testing CCP donors including Michelle Sexton, Haley Peden and Michael Pentella. We would also like to thank Julie Kurt and Gail Drey for assistance with our CCP program.

